# Unmet needs of Polish patients with cystic fibrosis seen through the lens of crowdfunding

**DOI:** 10.64898/2026.11.20.26368008

**Authors:** Mikołaj Kamiński, Jan K. Nowak, Zuzanna Karp, Aleksandra Borys, Jarosław Walkowiak

## Abstract

**Background:** Medical crowdfunding allows for funding for needs not covered by the local healthcare system. We aimed to characterize the crowdfunding campaigns for the needs of Polish individuals with cystic fibrosis (CF).

**Methods:** We analyzed data from a leading medical crowdfunding webpage in Poland, focusing on campaigns for individual medical needs initiated between 2009 and 2017. We characterized the features of individuals with CF and the declared goals for which the funds from the online campaigns were intended.

**Results:** Of all 2,656 campaigns, 96 (3.6%) were related to individuals with CF. Approximately two-thirds of these campaigns reached their target amount. Individuals with CF primarily raised funds to cover treatment costs, medical equipment, and entertainment. The successful collection had a significantly lower financial target than unsuccessful campaigns. If the receivers were living in a city or fundraising to gather funds for an inhaler, these were also associated with the campaign’s success. However, these associations did not reach statistical significance.

**Conclusions:** Crowdfunding campaigns can be an effective funding source for the needs of patients with CF that are not covered by the public healthcare system. Analyzing crowdfunding efforts for specific patient groups can help reveal their unmet health needs.

## Introduction

The costs of therapy are one of the most significant barriers to improving the quality of life of patients with cystic fibrosis (CF) [1]. The total yearly expense of the treatment of a patient with CF ranges between 23,300 and 68,700 euro [2]. Many public healthcare systems provide limited access to novel treatments for economic reasons. Therefore, people suffering from CF actively seek alternative methods of financing their needs. In our Department, most of the children with CF are supported by charity organizations (unpublished data).

Medical crowdfunding not only helps to meet health needs by pooling small contributions [3], but also increases rare disease awareness and might enable access to more advanced treatments. However, there are growing concerns that collected funds may be spent on unproven therapies. Medical crowdfunding is a poorly explored field of public health. We hypothesised that analysis of crowdfunding campaigns initiated for patients with CF may reveal both unmet health needs and an interest in unproven methods of treatment. This information underscores the urgent need to better educate CF patients on the most up-to-date evidence in order to help them establish reasonable aims.

We aimed to characterize the crowdfunding campaigns for the needs of Polish individuals with CF.

## Materials and methods

The research utilized publicly accessible data, and according to the regulations set forth by our University’s Bioethics Committee, we did not require approval. The analytical procedures were executed in full compliance with the platform’s established guidelines.

We analyzed data from the leading medical crowdfunding webpage in Poland, www.siepomaga.pl, managed by Siepomaga Foundation (Poznań, Poland), which declared that all receivers had been verified. We included the campaigns that started in 2009-2017 and finished before July 14th, 2018. We characterized features of the receiver, including the declared type of disease, age, category, sex, and place of living (urban area or not), the financial features of the collection, outcome (successful or not), number of Facebook shares, and duration of the campaign. The declared aims of the collection could be multiple. The use of the phrase “to make a patient’s dream come true” refers to entertainment purposes, e.g., toys or a holiday tour. A detailed description of the data collection process can be found in a recent report [4].

We conducted descriptive analyses. The data are presented as numbers (percentages) or median (25th–75th percentile). The Mann-Whitney U and Chi-square tests were used to compare successful vs. unsuccessful collections for individuals with CF. The cut-off for the p-value was set < 0.05. The dataset is provided as Supplementary File 1.

## Results

Among all the collections from 2009-2017 (n = 2,656), we identified 96 that aimed to support individuals with CF (3.6%). The receivers represented approximately 4.0% of people with CF in Poland [5]. Of 96 campaigns, 60 reached their financial target (62.5%). The most frequent comorbidities were: post-lung transplant condition n = 5 (5.2%), pancreatic insufficiency n = 4 (4.2%), and diabetes n = 4 (4.2%). The characteristics of the campaign and comparison of successful and unsuccessful collections are presented in Table 1. The successful collection had a significantly lower financial target than unsuccessful campaigns. Additionally, campaigns involving city residents or fundraising for an inhaler showed a trend towards greater success. However, these associations did not reach statistical significance. During data collection, a small percentage of campaigners specified the exact type of the medications (6 of 36). Therefore, we did not conduct a detailed analysis of the patient’s medication needs. Further details can be found in Supplementary File 1. Several examples of collection aims are provided in Supplementary Table 1.

**Table 1.** Comparison of successful and unsuccessful collections for needs of individuals with cystic fibrosis. Data presented as median (25^th^ percentile-75^th^ percentile) / n(%).

| Variable | All collections<br>n = 96 (100.0%) | Successful collections<br>n = 60 (62.5%) | Unsuccessful collections<br>n = 36 (37.5%) | p-value |
|---|---|---|---|---|
| **Target of the collection [EUR]** | **3,352 (1,234-6,028)** | **2,539 (884-4,657)** | **5,879 (3,251-11,146)** | **< 0.001** |
| Total raised sum [EUR] | 2,258 (9,25-4,267) | 2,540 (885-4,761) | 1,966 (1,027-3,845) | 0.62 |
| Total number of payments | 279 (96-612) | 303 (81-628) | 264 (114-553) | 0.99 |
| Mean payment values [EUR] | 7.87 (5.41-11.51) | 7.68 (5.75-13.31) | 7.99 (4.44-9.38) | 0.21 |
| Number of Facebook shares | 161 (0-869) | 227 (0-939) | 65 (0-554) | 0.47 |
| **Duration of the collection [days]** | **91 (15-235)** | **57 (11-111)** | **188 (93-263)** | **< 0.001** |
| Receiver lives in the city | 67 (70.0) | 46 (76.7) | 21 (58.3) | 0.06 |
| Sex: males | 48 (50.0) | 30 (50.0) | 18 (50.0) | 0.98 |
| Collection for at least two children | 6 (6.3) | 3 (5.0) | 3 (8.3) | 0.51 |
| Age: Child | 59 (61.0) | 39 (65.0) | 20 (55.6) | 0.36 |
| At least two different medical conditions declared | 14 (14.6) | 8 (13.3) | 6 (16.7) | 0.65 |
| Aim: uncovered treatment | 36 (37.5) | 21 (35.0) | 15 (41.7) | 0.51 |
| Aim: airway clearance vests | 14 (14.6) | 8 (13.3) | 6 (16.7) | 0.65 |
| Aim: dream come true | 13 (13.5) | 9 (15.0) | 4 (11.1) | 0.59 |
| Aim: costs associated with lung transplantation | 11 (11.5) | 6 (10.0) | 5 (13.9) | 0.56 |
| Aim: rehabilitation | 10 (10.4) | 6 (10.0) | 4 (11.1) | 0.86 |
| Aim: mobile oxygen concentrator | 10 (10.4) | 5 (8.3) | 5 (13.9) | 0.39 |
| Aim: inhaler | 6 (6.3) | 6 (10.0) | 0 (0.0) | 0.05 |

EUR - Euro

## Discussion

To the best of our knowledge, this is the first study to investigate medical crowdfunding for the needs of individuals with CF. The majority of fundraising campaigns for patients with CF involved children, accounting for 61.0% of cases. Overall, 70.0% of all campaign receivers lived in urban areas.

Interestingly, receivers residing in cities numerically achieved the target fundraising amount more often than those living outside urban areas. This could be associated with the fact that city dwellers are better at crafting compelling fundraising descriptions, or they might have larger social networks and make more effective use of online tools compared to those living in rural areas. However, in the main study, the origin of the receiver was not associated with whether the campaign would be successfully completed or not [4].

It seems logical that the smaller the target amount, the easier it is to raise it through crowdfunding. This might also explain why, when inhalers were the fundraising goals, there was a numerical association with a greater likelihood of increasing the success of the campaign.

The most common objective of these campaigns was to cover the costs of pharmacological treatment, which represented 37.5% of the fundraisers. We suspect this was due to the limited reimbursement for many drugs used in CF treatment. For example, cystic fibrosis transmembrane conductance regulator (CFTR) modulators have only been reimbursed for CF patients in Poland since March 2022 [6].

Interestingly, alongside the costs of medical treatment, equipment, or rehabilitation, approximately one in seven campaigners aimed to fund entertainment. While this might seem controversial, it is understandable that funding toys or electronic equipment may improve the quality of life of those who are seriously and chronically ill.

Our study has several limitations. Primarily, the data were sourced from a Polish portal that facilitates crowdfunding campaigns. Consequently, the conclusions may not be generalizable to patients with CF living outside of Poland. Secondly, the analysis of crowdfunding campaigns is limited by the data provided by users. Crowdfunding platforms typically publish only selected variables. That said, for specifics such as the names of particular drugs or drug classes to be funded by the campaign, one must rely on campaign photos or descriptions, and often many details are not disclosed. Nevertheless, the approach we presented could also be applied in other contexts, uncovering the needs reported by patients with CF and initiating fundraising campaigns on crowdfunding platforms.

## Conclusions

- Crowdfunding campaigns can be an effective source of funding for the needs of patients with CF that are not covered by the public healthcare system.
- Analyzing crowdfunding efforts for specific patient groups can help reveal their unmet health needs.

## Data Availability

The dataset is included in the Supplementary File 1.

## Acknowledgements

Not applicable.

## Statement of competing interests

The authors declare no conflicts of interest.

## List of abbreviations

CF: cystic fibrosis
CFTR: cystic fibrosis transmembrane conductance regulator

## Notes

### Competing Interest Statement

The authors have declared no competing interest.

