## Supplementary Table 1 for "Unmet needs of Polish patients with cystic fibrosis seen through the lens of crowdfunding"

Examples of successful collections for needs of patients with cystic fibrosis (www.siepomaga.pl).

| **Characteristic** | **Declared aim** | **Facebook shares** | **Collected sum (EUR)** |
| --- | --- | --- | --- |
| Female, 11–15 years | costs of post-transplantation immunosuppression | 1,417 | 35,542 |
| Female, 6–10 years | airway clearance vests | 2,026 | 8,068 |
| Female, 21–25 years | mobile oxygen concentrator | 860 | 3,949 |
| Male, 11–15 years | costs of one-year rehabilitation | 243 | 1,730 |
| Female, 16–20 years | digital camera | 590 | 572 |

*Note: Patient ages are reported in non-overlapping 5-year ranges (0–5, 6–10, 11–15, 16–20, 21–25 years, etc.) to protect participant anonymity.*
